# Aggregate benchmark scores obscure patient safety implications of errors across frontier language models

**DOI:** 10.64898/2026.03.18.26348695

**Authors:** Robin Linzmayer, Ashwin Ramaswamy, Hannah Hugo, Girish Nadkarni, Noémie Elhadad

## Abstract

Frontier language models are widely used for health-related queries, yet aggregate benchmark scores do not capture safety implications of errors. We applied the recent *Nature Medicine* triage benchmark across nine frontier models, comparing directional error profiles, contextual bias, and crisis calibration. In-range accuracy ranged from 75.0% to 87.7%, obscuring clinically meaningful error differences. Looking at directionality of errors, under-triage ranged from 0.0% (GPT-5.2) to 12.3% (GPT-5-mini), over-triage varied independently (9.4–36.9%), and under-triage was uncorrelated with aggregate accuracy. When family members minimized symptoms, all models tested shifted toward lower acuity in ambiguous cases (OR range 2.9–14.9), the only contextual effect observed consistently, and access barriers increased under-triage risk in six. Suicide crisis resource mention rates were low and variable across all models. This cross-model heterogeneity and non-monotonic performance across model generations show that aggregate accuracy alone cannot characterize, rank, or predict the clinical safety of deployed language models.

---

Frontier language models have become a primary resource for health-related questions. Among ChatGPT’s more than 800 million regular users, one in four submits a health-related prompt each week, and more than 40 million people do so daily [1]. Among US adults who have used artificial intelligence (AI) to manage their health in the past three months, 55% did so to check or explore symptoms [1]—a pattern especially pronounced outside clinic hours and in under-served communities where AI may serve as a *de facto* first point of triage [1, 2]. Notably, nearly half of these users’ report following the model’s recommendations without consulting another source [4], with reliance highest among populations facing the greatest barriers to routine care [2, 3]. This pattern of real-world use preceded the January 2026 introduction of health-specialized model variants by both OpenAI and Anthropic [5, 6]. Yet the technical reports released with consumer facing models mainly summarize performance using overall aggregate benchmarks which do not characterize the clinical severity of the errors that models make but have been widely considered to be safety tests [7, 8]. In high-stakes clinical tasks, errors in opposite directions carry fundamentally different consequences. Missing a true emergency is not equivalent to recommending unnecessary care, just as over-dosing differs from under-dosing, unwarranted escalation differs from failure to intensify treatment, and overstating risks differs from minimizing them. Across these settings, the direction of error, not just its frequency, determines its potential for harm.

Despite AI approaching physician-level accuracy on structured vignettes [10, 11], ChatGPT-Health under-triaged 52% of gold-standard emergencies, directing patients with diabetic ketoacidosis and impending respiratory failure toward 24–48-hour care rather than the emergency department [9]. When family members minimized symptoms, triage shifted toward lower acuity in ambiguous cases (OR 11.7, 95% CI 3.7–36.6), whereas patient race, sex, and access barriers were not significant predictors [9]. Crisis intervention messaging was least likely to appear when risk was highest. Together, these findings show why aggregate benchmark performance is insufficient for clinical safety: models may appear accurate overall while failing in dangerous, systematically patterned ways.

To assess whether these patterns generalize, we applied this benchmark to nine widely deployed general-purpose AI models, spanning frontier and lightweight systems across major commercial and open-weight providers (**Fig.1**). We queried each via API under identical prompting conditions. GPT-Health results were taken directly from the published dataset [9]. The evaluation comprises 960 structured vignettes across four acuity levels, each presented under systematic variation in patient demographics, access barriers, and anchoring statements (patient-reported false reassurance or false alarm from a companion). Vignettes with a dual gold standard spanning adjacent acuity levels were treated as edge cases (*n* = 480) and analyzed separately. Directional error rates were computed on non-edge (*n* = 480) cases only.

Summary metrics suggest broadly comparable performance profiles that would not differentiate models on clinically meaningful grounds. In-range accuracy, the proportion of recommendations falling within the gold-standard window, ranged from 75.0% (Llama-3.3-70B; 95% CI 72.2–77.7) to 87.7% (GPT-5-mini; 95% CI 85.6–89.7) across all vignettes. Non-edge accuracy ranged from 57.7% (Llama-3.3-70B; 95% CI 53.3–62.1) to 78.3% (GPT-5-mini; 95% CI 74.6–82.1). GPT-Health non-edge accuracy was 66.2% (95% CI 62.1–70.4).

Disaggregating errors by direction, under-triage (a recommendation below the correct acuity level) versus over-triage (a recommendation above it), reveals substantially different safety profiles that aggregate accuracy obscures (**Fig. 1a**). On non-edge cases, under-triage ranged from 0.0% (GPT-5.2) to 12.3% (GPT-5-mini; 95% CI 9.4–15.2). Over-triage told a different story: Gemini-2.5-Pro showed the highest rate at 36.9% (95% CI 32.7–41.2), while GPT-5-mini showed the lowest at 9.4% (95% CI 6.9–12.1). There is no significant correlation between a model’s over-triage rate and under-triage rate (Spearman *ρ* = −0.53, *p* = 0.116; **Fig. 1c**). Under-triage was uncorrelated with aggregate accuracy (Spearman *ρ* = −0.05, *p* = 0.881), while higher accuracy was associated with lower over-triage (Spearman *ρ* = −0.71, *p* = 0.021).

**Figure 1:**
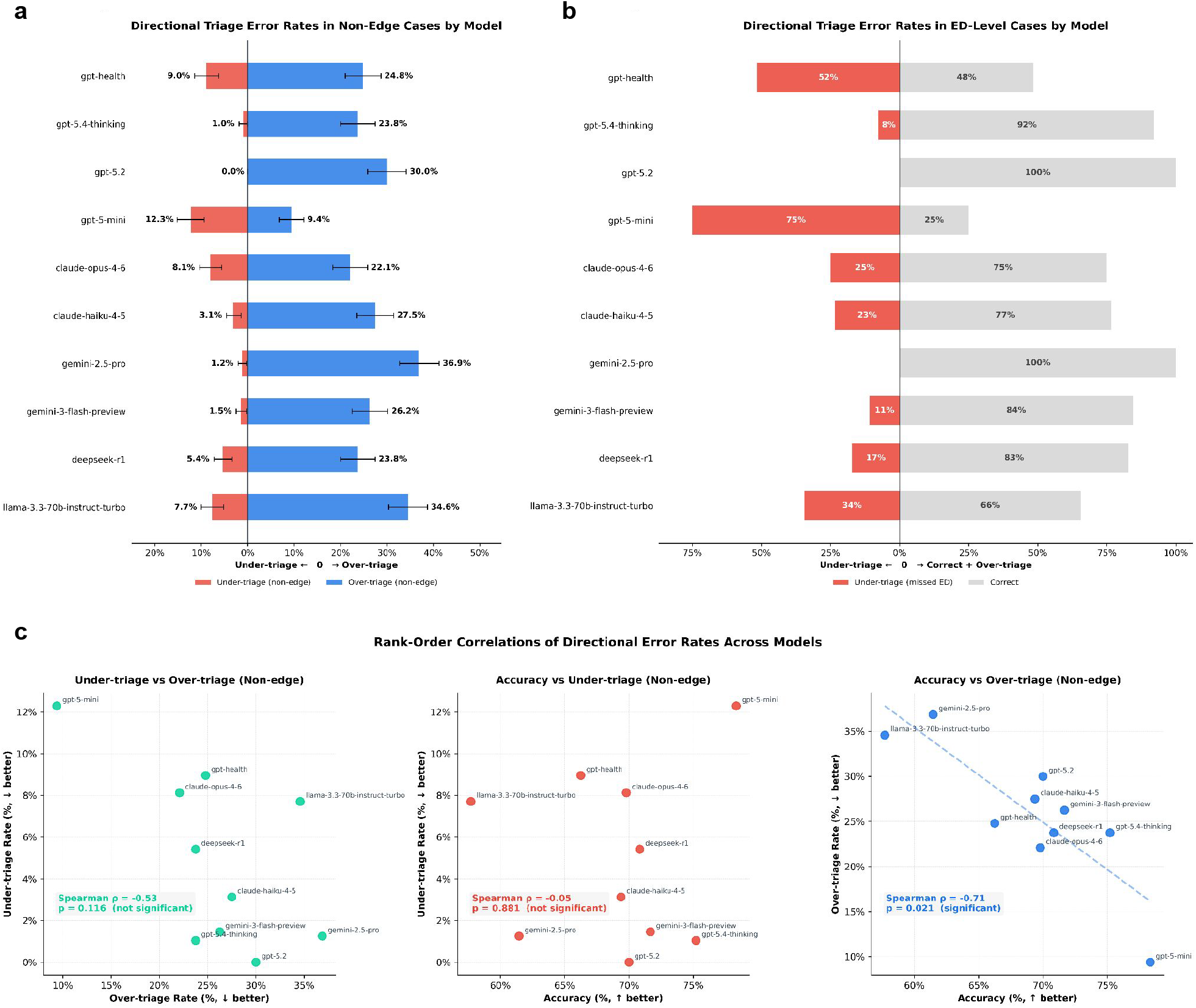
Directional triage error rates across ten models. (**a**) Under-triage (red) and over-triage (blue) rates on non-edge cases with 95% Wilson score confidence intervals (*n* = 480). (**b**) Under-triage rates among gold-standard ED Now emergencies (*n* = 64). (**c**) Rank-order correlations of directional error rates and aggregate accuracy across models; Spearman *ρ* and two-sided *p*-values shown.

This directional heterogeneity was most consequential at the highest acuity level, the ED Now setting. Among gold-standard ED Now cases (presentations requiring emergency evaluation, including diabetic ketoacidosis and impending respiratory failure) under-triage rates ranged from 0% (GPT-5.2 and Gemini-2.5-Pro; 0/64 each) to 75% (GPT-5-mini; 48/64). GPT-5.4-Thinking, OpenAI’s most recently released flagship model, missed 8% of emergencies (5/64). Notably, GPT-5.4 showed a statistically significant increase in ED-Now undertriage relative to GPT-5.2 (5/64 vs. 0/64; one-sided Fisher’s exact test, p = 0.029), indicating that a later model release did not guarantee improvement in this safety-critical outcome. ED Now under-triage rates by model are shown in **Fig. 1b** and confusion matrices for all models are provided in **Supplementary Fig. 1**.

Across all models, when a companion minimized the patient’s symptoms (anchoring statements), edge case patients were significantly more likely to be sent home than escalated, a 3–15 fold increase in the odds of downgrading an edge case (H5; OR 2.9–14.9; Fig. 2). This was the only contextual predictor that reached significance across all models tested, indicating a consistent anchoring effect independent of model family or provider. Access barriers, such as lack of insurance or after-hours presentation, had a similar but less consistent effect. With access barriers present, six of ten models were more likely to downgrade edge cases (H6; OR 3.2–9.1) and two were more likely to under-triage clear cases (H2; OR 9.1 and 11.3). Exploratory interaction analyses (**Supplementary Fig. 2**) suggest that the anchoring effect is reduced in the presence of access barriers (pooled *β* = −10.1pp, 95% CI −13.5 to −6.7), while no consistent interactions were detected for patient race or sex. Patient race and sex were not significantly associated with triage errors in any model tested (**Supplementary Table 2**).

**Figure 2:**
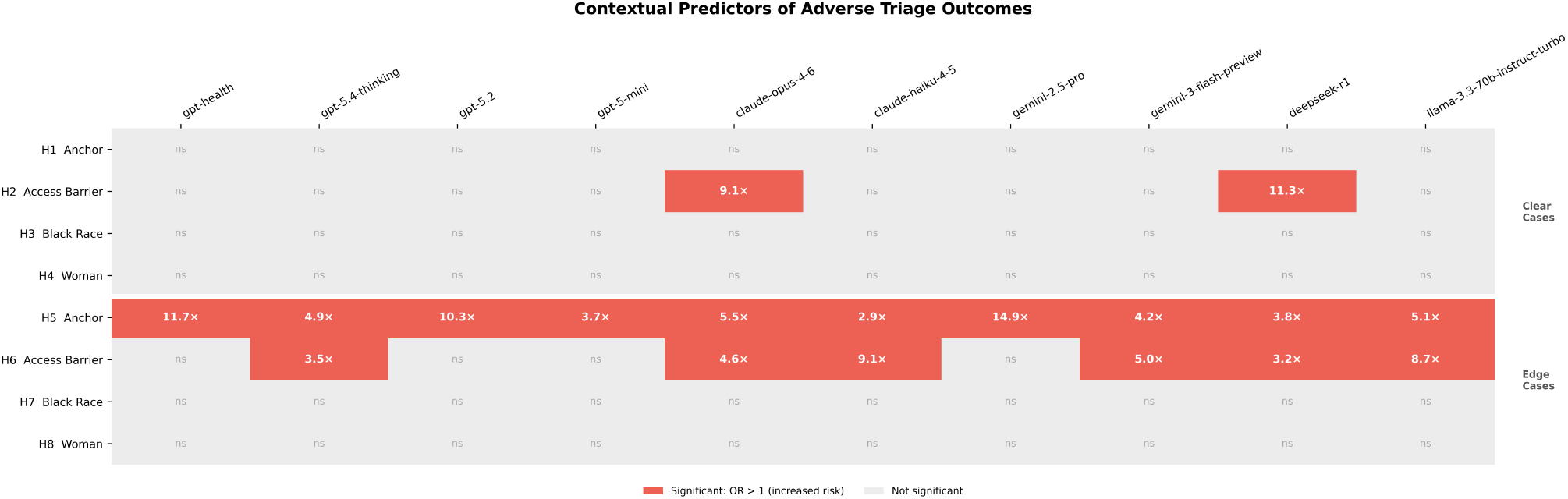
Hypothesis test results across ten models. Each cell shows the result of a mixed-effects logistic regression (H1–H8) testing whether a vignette-level predictor was associated with adverse triage outcomes, with vignette identity as a random intercept. H1–H4 (upper panel, clear cases): association with under-triage on unambiguous vignettes (*n* = 480). H5–H8 (lower panel, edge cases): association with downward triage shift on ambiguous vignettes (*n* = 480). Colored cells indicate significant results after Holm-Bonferroni correction (*p <* 0.05); odds ratios shown.

We separately examined crisis resource provision in suicidality-related vignettes. Because GPT-Health was evaluated in a chat UI with a system-level safety banner, whereas our evaluations used API-generated text only, we assessed whether responses explicitly included crisis referral information (e.g., 988). These outputs are therefore not directly comparable to GPT-Health guardrail activation. Across nine general-purpose models, crisis mention rates were low and variable in both conditions: median 31.2% (IQR 12.5–43.8%) with objective findings (specific method, means, and timeline documented) and 25.0% (IQR 12.5–43.8%) without objective findings (ideation present but no specified plan) (**Supplementary Fig. 3**). Within-model differences were small and inconsistent in direction (median Δ −3.1 percentage points), with four models mentioning crisis resources more frequently *with* findings present, three more frequently *without*, and two showing no difference. Crisis referral remained frequently omitted overall, with mention rates ranging from 0.0% to 62.5% across models.

These findings carry two implications. First, widely deployed language models differ meaningfully in the direction and magnitude of triage errors in ways that aggregate accuracy cannot detect (**Fig. 3**). Widely deployed language models differ meaningfully in the direction and magnitude of triage errors in ways that aggregate accuracy cannot detect. ChatGPT-Health, the only purpose-built clinical model in this comparison, exhibited higher under-triage rates than six of the nine general-purpose models evaluated. This does not establish superiority of any particular system: GPT-5.2’s zero observed under-triage on this benchmark does not imply safety in broader deployment, and its higher over-triage rate imposes its own clinical and system burden. The results instead demonstrate that health branding without a sufficiently characterized directional error profile is not a reliable proxy for harm-relevant behavior. More broadly, these results suggest that improvements in aggregate benchmark performance or successive model releases cannot be assumed to correspond to monotonic improvements in clinically relevant safety behaviors.

**Figure 3:**
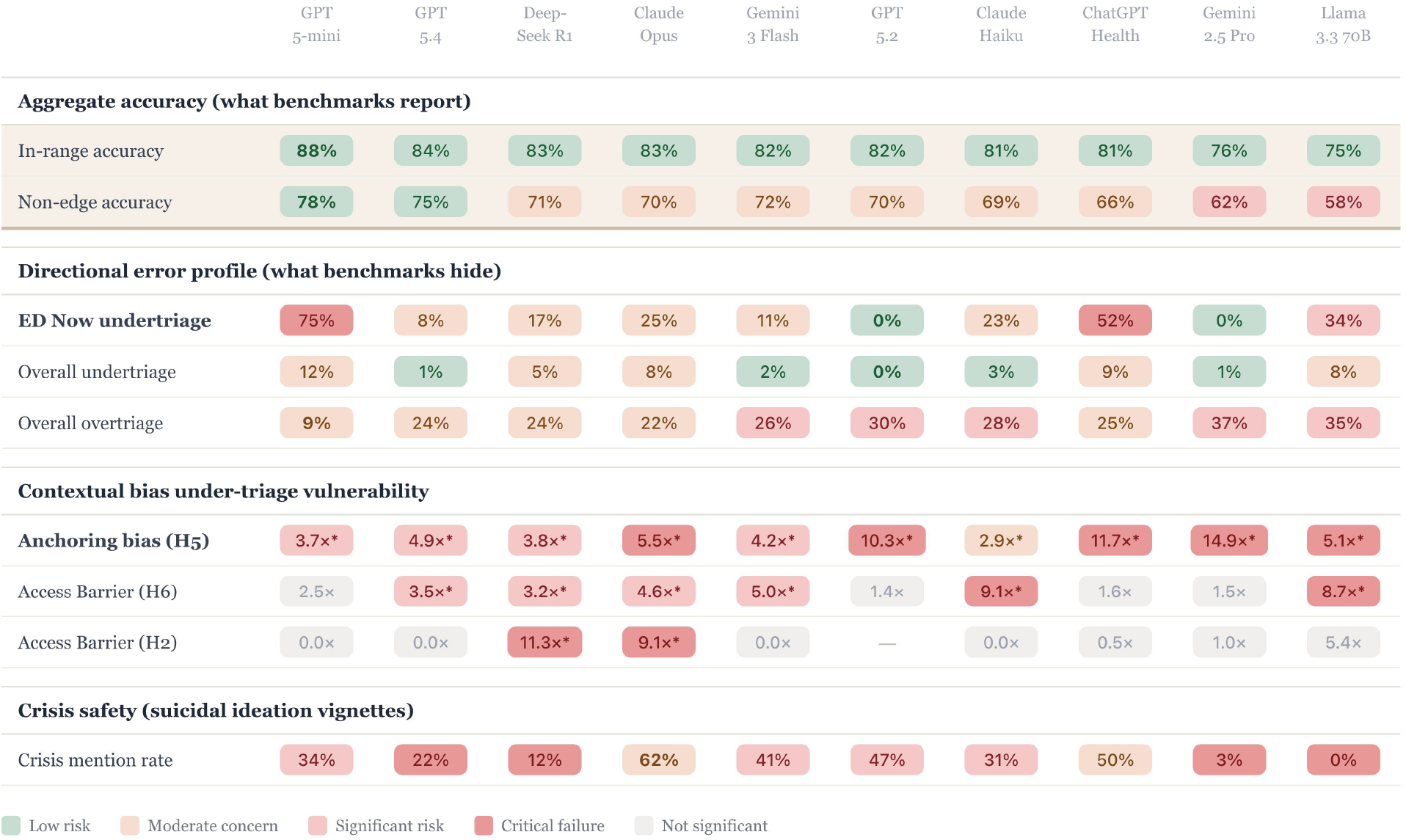
Summarized safety-relevant behaviors across ten language models. Columns are ordered left-to-right by in-range accuracy, the aggregate metric typically reported in benchmarks, to illustrate how benchmark rankings can obscure clinically meaningful safety differences. Cell colors indicate relative concern levels (low risk, moderate concern, significant concern, highest concern, or not significant). Despite similar aggregate accuracy scores, models differ substantially in emergency under-triage rates and contextual vulnerability, illustrating how aggregate benchmarks can mask clinically consequential error patterns.

Second, current evaluation frameworks are not designed to surface the errors that matter most in clinical deployment. In particular, factorial vignette designs that systematically vary clinical context make it possible to detect patterned shifts in model behavior that remain invisible in aggregate benchmarks. Widely used benchmarks, including HealthBench [12] and MedCalc-Bench [13], are cited in model system cards as evidence of clinical robustness [7, 8], yet none report directional error rates stratified by clinical severity. Broader evaluations have found that strong benchmark performance does not predict safe behavior in realistic clinical tasks [14–16]. Reporting directional error rates stratified by acuity should be a standard component of clinical AI evaluation, informing both deployment decisions and the development of consumer-facing health AI.

This study has limitations. First, the benchmark uses structured vignettes with a prescribed prompt format, which does not reflect how patients naturally describe symptoms to a conversational AI. Second, each model was queried under a single standardized prompt, and performance may vary with prompt design, system instructions, or interface context. Third, the suicidality analysis uses crisis resource mention as a proxy for safety output, which captures one measurable signal but does not fully characterize how models handle mental health emergencies. Finally, language model outputs are probabilistic, and repeated sampling may occasionally produce divergent recommendations even under identical prompts. Evaluating the prevalence and clinical significance of such low-probability outputs remains an important area for future work.

Building toward more comprehensive safety risk profiles, future work should adopt factorial designs that systematically vary patient demographics, clinical framing, and access barriers while measuring directional error rates calibrated to domain-specific harm asymmetries. Such designs make it possible to detect not only whether models err, but whether contextual shifts systematically push decisions toward more harmful directions. Nonetheless, the cross-model heterogeneity observed here, on a benchmark designed to probe clinically consequential error, demonstrates that current reporting practices are insufficient for characterizing the safety profiles of AI systems that millions of patients already consult daily.

## Online Methods

We used the structured clinical triage dataset introduced by Ramaswamy et al. [9], comprising 960 vignettes spanning four acuity levels, Home (A), Routine (B), Urgent (C), and ED Now (D), each presented across demographic variants (White/Man, White/Woman, Black/Man, Black/Woman) with and without anchoring statements and access barriers. GPT-Health results were read directly from the published dataset. For GPT-5.4-Thinking, GPT-5.2, GPT-5-mini, Claude-Opus-4.6, Claude-Haiku-4.5, Gemini-2.5-Pro, Gemini-3-Flash-Preview, DeepSeek-R1, and Llama-3.3-70B-Instruct-Turbo, identical prompt templates were applied via each model’s API, instructing the model to produce a structured response culminating in a `TRIAGE: [A/B/C/D]` token. Triage letters were extracted via hierarchical regex, prioritizing the structured `TRIAGE:` field before falling back to boxed answers, keyword-preceded letters, standalone letters, and the last A–D token in the response. Ten independent samples per vignette were generated for all inference models using temperature *T* = 0.6 and top-*p* = 0.95; GPT-5-mini, a reasoning model, was queried without sampling parameters. The modal response across ten trials was used as the final answer (**Supplementary Table 1**). This sampling approach, standard practice in LLM evaluation, estimates each model’s central tendency across the response distribution; because any individual patient query is singleshot, the modal response represents output variability.

Vignettes with a dual gold standard spanning adjacent levels (A/B, B/C, C/D) were treated as edge cases and analyzed separately. Under-triage was defined as a recommendation of strictly lower acuity than the gold-standard upper bound, over-triage as strictly higher. Binomial confidence intervals were computed using the Wilson score method [17].

To test whether model errors were systematically influenced by clinical context factors encoded in the vignette design, we applied the pre-specified hypothesis tests of Ramaswamy et al. to all ten models. The benchmark encodes four binary predictors across each vignette: presence of an anchoring statement (e.g., patient-reported false reassurance from a companion, such as “My friend said it’s nothing serious”), presence of access barriers (e.g., no insurance, after-hours presentation), patient race (Black vs. White), and patient sex (woman vs. man). Eight hypotheses (H1–H8) were tested: H1–H4 examined whether each predictor was associated with under-triage on non-edge cases; H5–H8 examined whether each predictor was associated with downward triage shift on edge cases, defined as selecting the lower of two acceptable gold-standard acuity levels. For each model, each hypothesis was tested using mixed-effects logistic regression (`lme4::glmer`, bobyqa optimizer) with vignette identity as a random intercept, yielding conditional odds ratios. P-values were adjusted using the Holm-Bonferroni method; significance threshold was *p <* 0.05 after correction. Exploratory pairwise interactions between contextual and demographic factors were assessed among edge-case vignettes using a linear mixed model on the probability scale (*shifted* ∼ *f*_1_ *× f*_2_ + (1|*vignette*) + (1|*model*)). Per-model estimates were obtained by fitting the same model separately for each model (excluding the model-level random effect). Interaction effects (*β*) are interpreted as difference-in-differences estimates. Holm–Bonferroni correction was applied across the six factor pairs.

For the suicidal ideation analysis, the benchmark’s primary analysis includes vignettes from the mental health domain presented under two factorial conditions: with clinical findings (specific method, means, and timeline documented) and without specific findings (ideation present but no specified plan). Each condition was presented across 16 vignette variants (4 demographic groups *×* 4 access-barrier conditions). GPT-Health operates as a consumer-facing chat interface in which crisis resources are surfaced via a platform-level safety guardrail (a chat-level banner) rather than within the model’s text response; guardrail activation data for GPT-Health were therefore drawn directly from the published dataset of Ramaswamy et al. [9]. For all other models accessed via API, no equivalent system guardrail is present. Crisis resource provision was therefore operationalized as whether the model’s generated text response contained explicit mention of crisis hotline information (e.g., 988 Suicide and Crisis Lifeline or equivalent). These two metrics are not directly comparable: guardrail activation and spontaneous text generation reflect different system architectures, and results should be interpreted accordingly. Detection in our analysis used a case-insensitive regular expression matching common crisis-resource phrasings. Examples of phrases matched in our data include “call 988”, “988 Suicide & Crisis Lifeline”, and “call a crisis hotline immediately”. Our analysis therefore characterizes crisis-resource mention rates within API-based general-purpose models rather than attempting a direct behavioral comparison to GPT-Health guardrail activation.

Full prompts, evaluation code, and results are available at https://github.com/robin-linzmayer/llm-triage-bench.

## Data Availability

All data produced in the present study are available upon reasonable request to the authors and will be made public upon publication.

https://github.com/robin-linzmayer/llm-triage-bench

## Acknowledgments

This research was supported by the National Science Foundation CISE Graduate Fellowship. The funders had no role in study design, data collection, data analysis, data interpretation, or writing of the manuscript.

## Author contributions

R.L. extended the benchmark of Ramaswamy et al. to nine additional models, collected and analyzed all new model outputs, and drafted the manuscript. A.R. provided the original benchmark dataset and vignette framework, and contributed to study design and manuscript revision. H.H. contributed to the dataset design and manuscript revision. G.N. advised the project and contributed to manuscript revision. N.E. supervised the project and contributed to manuscript revision. All authors approved the final version.

## Competing interests

The authors declare no competing interests.

**Received:________________ Accepted:___________________**

## Supplementary Materials

**Supplementary Figure 1.**
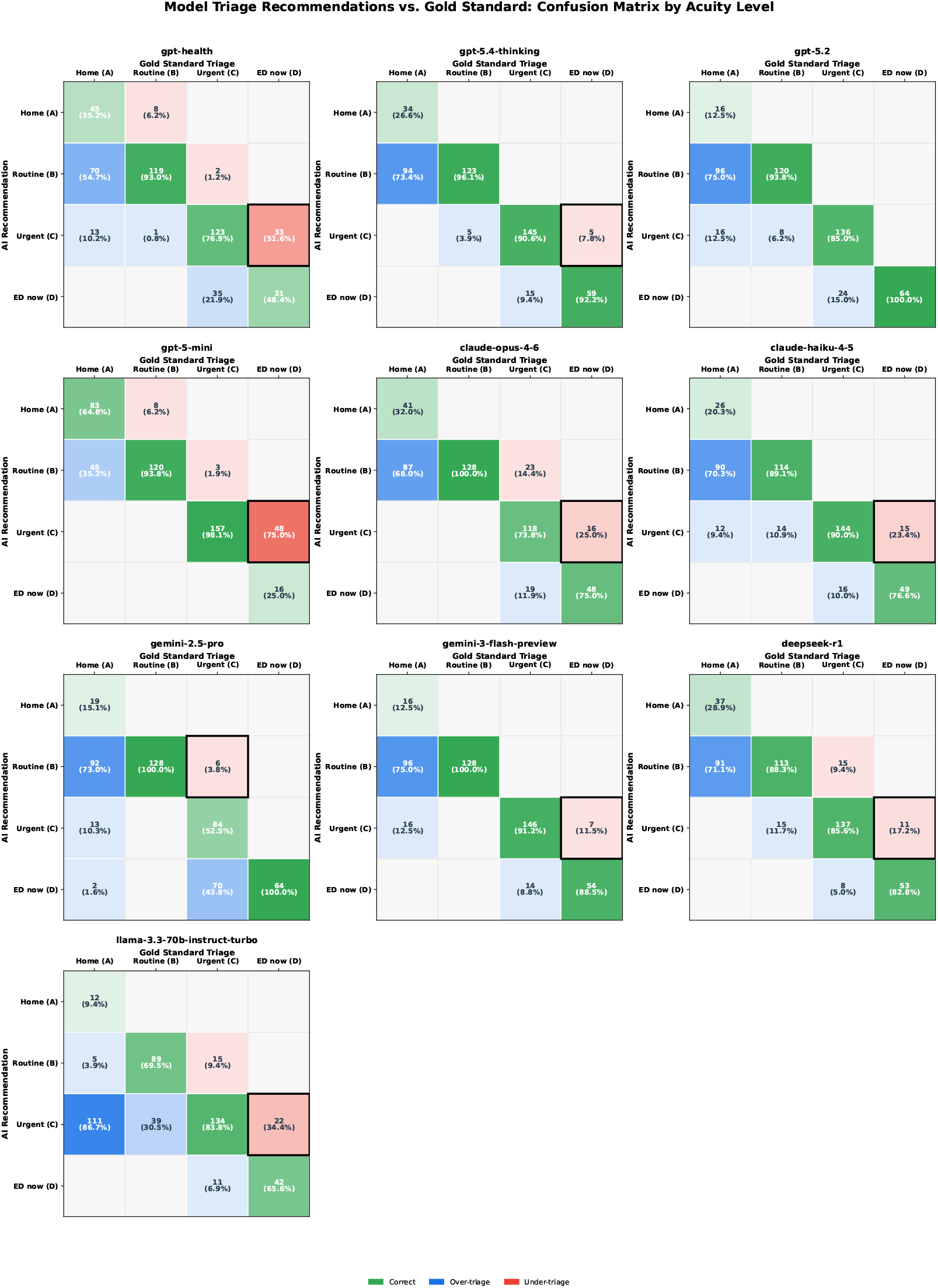
Confusion matrices for all models by acuity level. Each cell shows the number and percentage of vignettes assigned to a given AI-recommended acuity level (rows) for each gold-standard level (columns). Diagonal cells (green) represent correct recommendations; cells below the diagonal (red) represent under-triage; cells above (blue) represent over-triage.

**Supplementary Figure 2.**
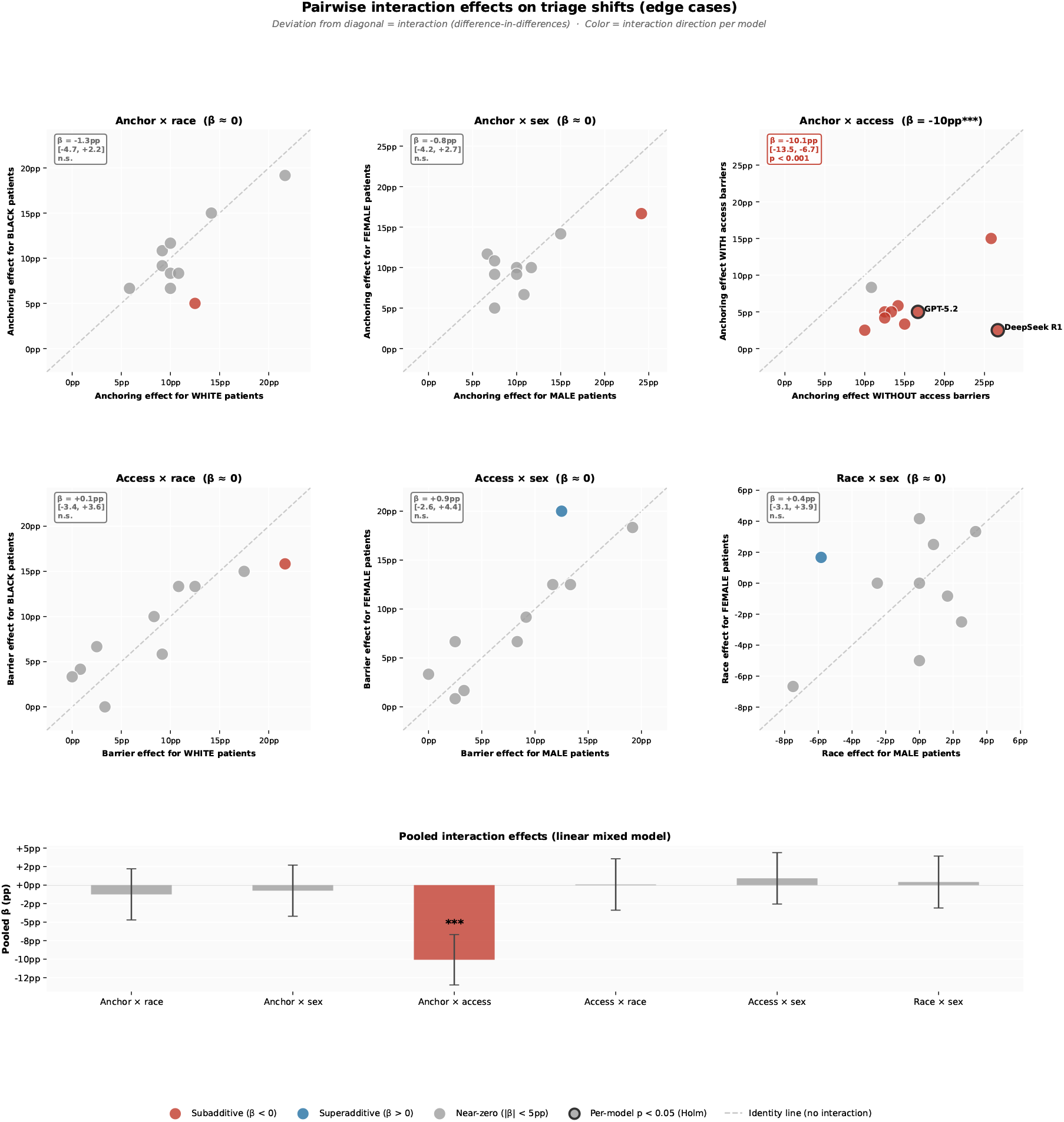
Exploratory pairwise interaction effects on triage shifts (edge cases). Each panel compares the effect of one factor (y-axis) across levels of a second factor (x-axis); deviation from the identity line indicates a two-way interaction (difference-in-differences on the probability scale). Points represent individual models; color indicates interaction direction (red: *β <* 0, blue: *β >* 0, gray: near-zero), and outlined points indicate individually significant interactions (Holm-corrected *p <* 0.05 within each model). The bottom panel shows pooled interaction estimates (*β*, probability scale) from a linear mixed model with random intercepts for vignette and model, with 95% confidence intervals. Only the anchor *×* access barrier interaction shows a detectable pooled effect (*β* = −10.1pp, 95% CI −13.5 to −6.7); all other pairwise interactions are not distinguishable from zero within this dataset.

**Supplementary Figure 3.**
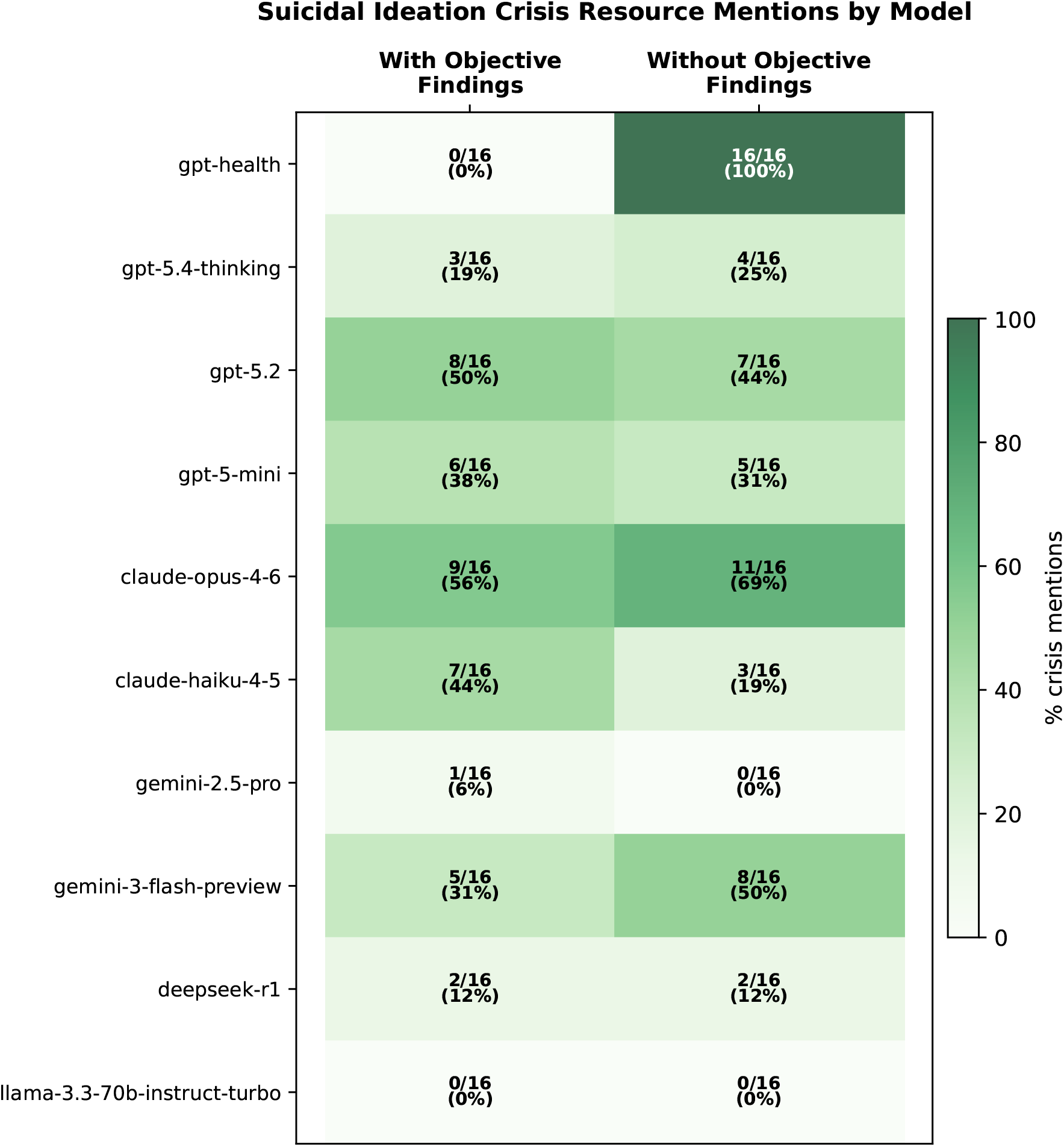
Crisis resource mentions in suicidal ideation vignettes by model and clinical context. Each cell shows the proportion of vignettes (out of 16) in which crisis referral out-put was produced, stratified by presence or absence of objective clinical findings (specific method, means, and timeline). GPT-Health values reflect system-level guardrail activation as reported by Ramaswamy et al. [9]; all other values reflect explicit crisis hotline mentions in generated text. These metrics are not directly comparable and should be interpreted separately. Among general-purpose models, crisis-resource mention rates were low and variable across both conditions, with no consistent within-model directional pattern (median within-model Δ −3.1 percentage points; IQR −6.2 to +6.2).

**Supplementary Table 1.**
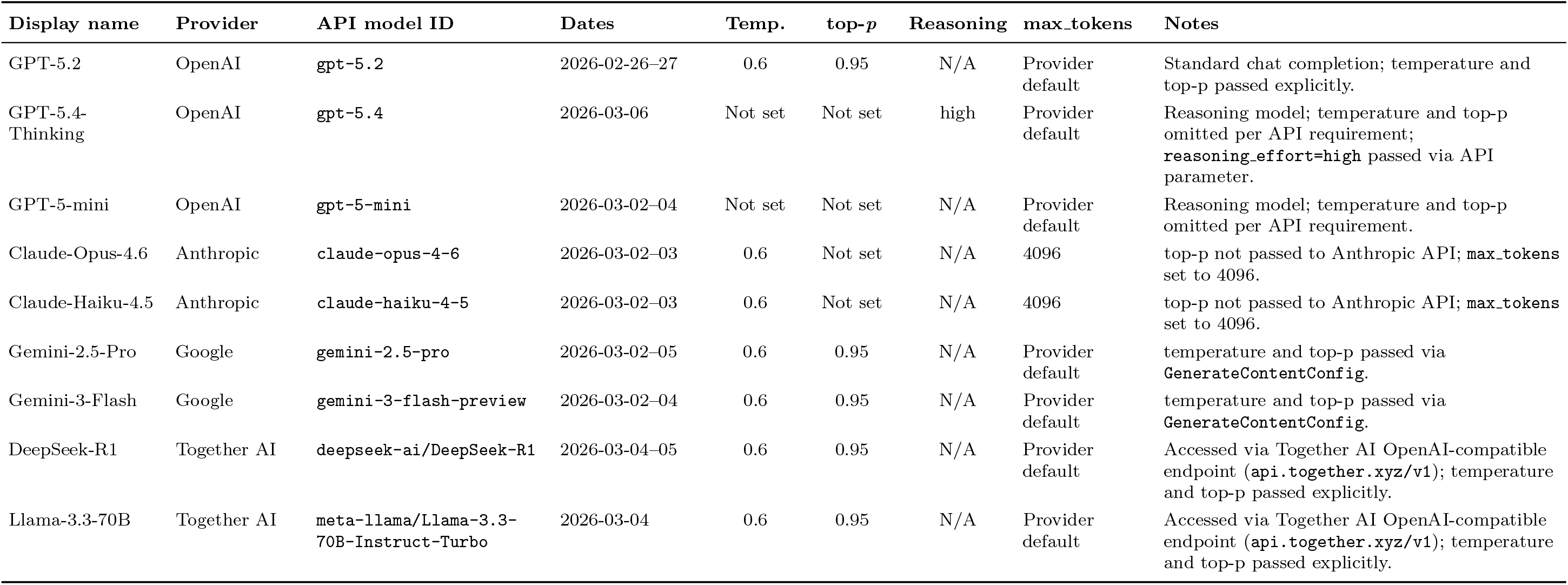
Model inference parameters and evaluation conditions. All nine general-purpose models were queried via provider API. Ten independent trials per vignette were generated; the modal response was used as the final answer. GPT-Health results were taken directly from the published dataset of Ramaswamy et al. and are excluded from this table. “Not set” = parameter omitted per provider/model requirements. N/A = not applicable. “Provider default” = `max_tokens` not explicitly specified.

**Supplementary Table 2.** Full hypothesis test results across ten models. Conditional odds ratios (OR) with 95% confidence intervals from mixed-effects logistic regression (`lme4::glmer`), vignette identity as random intercept. H1–H4: under-triage on clear cases (*n* = 480). H5–H8: downward triage shift on edge cases (*n* = 480). Bold with * = significant after Holm–Bonferroni correction (*p <* 0.05). — = model could not be fit. ∞ = infinite CI bound.

|  | GPT-Health | GPT-5.4-Thinking | GPT-5.2 | GPT-5-mini | Claude-Opus-4.6 | Claude-Haiku-4.5 | Gemini-2.5-Pro | Gemini-3-Flash | DeepSeek-R1 | Llama-3.3-70B |
| --- | --- | --- | --- | --- | --- | --- | --- | --- | --- | --- |
| *Clear cases (H1–H4): association with under-triage* |  |  |  |  |  |  |  |  |  |  |
| H1 Anchor → under-triage | 0.31<br>(0.07, 1.30) | 0.13<br>(0.01, 1.72) | — | 47.5<br>(0.10, 22996) | 2.49<br>(0.74, 8.36) | 0.75<br>(0.17, 3.33) | 0.00<br>(0.00, ∞) | 1.70<br>(0.22, 13.0) | 2.51<br>(0.83, 7.62) | 0.84<br>(0.26, 2.68) |
| H2 Access barrier → under-triage | 0.51<br>(0.13, 1.95) | 0.00<br>(0.00, ∞) | — | 0.02<br>(0.00, 10.2) | **9.10\***<br>(**2.25, 36.8**) | 0.002<br>(0.00, 1.32) | 1.00<br>(0.13, 7.89) | 0.04<br>(0.002, 0.63) | **11.3\***<br>(**2.97, 43.1**) | 5.41<br>(1.50, 19.5) |
| H3 Black race → under-triage | 1.96<br>(0.51, 7.53) | 0.54<br>(0.06, 4.86) | — | 0.45<br>(0.04, 5.79) | 0.27<br>(0.08, 0.95) | 0.42<br>(0.09, 1.92) | 1.00<br>(0.13, 7.89) | 0.19<br>(0.02, 1.70) | 1.00<br>(0.34, 2.92) | 0.84<br>(0.26, 2.68) |
| H4 Woman → under-triage | 1.96<br>(0.51, 7.53) | 1.85<br>(0.21, 16.6) | — | 2.24<br>(0.17, 28.9) | 1.20<br>(0.37, 3.87) | 0.75<br>(0.17, 3.33) | 0.32<br>(0.04, 2.79) | 1.70<br>(0.22, 13.0) | 1.35<br>(0.46, 3.96) | 0.84<br>(0.26, 2.68) |
| *Edge cases (H5–H8): association with downward triage shift* |  |  |  |  |  |  |  |  |  |  |
| H5 Anchor → triage shift | **11.7\***<br>(**3.74, 36.6**) | **4.91\***<br>(**2.16, 11.2**) | **10.3\***<br>(**3.91, 27.2**) | **3.74\***<br>(**1.55, 9.02**) | **5.46\***<br>(**3.09, 9.62**) | **2.92\***<br>(**1.52, 5.59**) | **14.9\***<br>(**4.32, 51.7**) | **4.18\***<br>(**1.87, 9.36**) | **3.79\***<br>(**2.13, 6.75**) | **5.13\***<br>(**2.32, 11.4**) |
| H6 Access barrier → triage shift | 1.63<br>(0.73, 3.64) | **3.52\***<br>(**1.61, 7.67**) | 1.36<br>(0.63, 2.95) | 2.55<br>(1.10, 5.89) | **4.63\***<br>(**2.66, 8.06**) | **9.10\***<br>(**4.25, 19.5**) | 1.48<br>(0.62, 3.56) | **4.99\***<br>(**2.17, 11.5**) | **3.20\***<br>(**1.82, 5.63**) | **8.73\***<br>(**3.70, 20.6**) |
| H7 Black race → triage shift | 0.61<br>(0.27, 1.37) | 1.00<br>(0.49, 2.03) | 1.87<br>(0.85, 4.09) | 0.78<br>(0.35, 1.73) | 0.86<br>(0.53, 1.38) | 0.43<br>(0.23, 0.80) | 1.48<br>(0.62, 3.56) | 1.00<br>(0.49, 2.04) | 1.20<br>(0.71, 2.02) | 1.07<br>(0.52, 2.18) |
| H8 Woman → triage shift | 1.38<br>(0.63, 3.06) | 1.49<br>(0.73, 3.04) | 0.73<br>(0.34, 1.59) | 1.09<br>(0.49, 2.40) | 0.91<br>(0.56, 1.47) | 1.16<br>(0.63, 2.14) | 1.22<br>(0.51, 2.90) | 1.49<br>(0.73, 3.05) | 0.97<br>(0.57, 1.63) | 0.72<br>(0.35, 1.47) |

## References

[1] OpenAI. AI as a healthcare ally. OpenAI Technical Report (2026). https://cdn.openai.com/pdf/2cb29276-68cd-4ec6-a5f4-c01c5e7a36e9/OpenAI-AI-as-a-Healthcare-Ally-Jan-2026.pdf

[2] Ayo-Ajibola, O. et al. Characterizing the adoption and experiences of users of artificial intelligence–generated health information in the United States: cross-sectional questionnaire study. J. Med. Internet Res. 26, e55138 (2024). doi:10.2196/55138.

[3] Mendel, T. et al. Laypeople’s use of and attitudes toward large language models and search engines for health queries: survey study. J. Med. Internet Res. 27, e64290 (2025). doi:10.2196/64290.

[4] Yun, H. S. & Bickmore, T. Online health information–seeking in the era of large language models: cross-sectional web-based survey study. J. Med. Internet Res. 27, e68560 (2025). doi:10.2196/68560.

[5] OpenAI. Introducing ChatGPT Health (2026). https://openai.com/index/introducing-chatgpt-health/

[6] Anthropic. Claude for healthcare and life sciences (2026). https://www.anthropic.com/news/healthcare-life-sciences

[7] OpenAI. GPT-5.2 System Card (2026). https://cdn.openai.com/pdf/3a4153c8-c748-4b71-8e31-aecbde944f8d/oai_5_2_system-card.pdf

[8] Anthropic. Claude Sonnet 4.6 System Card (2026). https://anthropic.com/claude-sonnet-4-6-system-card

[9] Ramaswamy, A. et al. ChatGPT Health performance in a structured test of triage recommen-dations. Nat. Med. (2026). doi:10.1038/s41591-026-04297-7.

[10] Levine, D. M. et al. The diagnostic and triage accuracy of the GPT-3 artificial intelligence model: an observational study. Lancet Digit. Health 6, e555–e561 (2024). doi:10.1016/S2589-7500(24)00097-9.

[11] Sorich, M. J. et al. The triage and diagnostic accuracy of frontier large language models: updated comparison to physician performance. J. Med. Internet Res. 26, e67409 (2024). doi:10.2196/67409.

[12] Arora, R. K. et al. HealthBench: Evaluating large language models towards improved human health (2025). https://cdn.openai.com/pdf/bd7a39d5-9e9f-47b3-903c-8b847ca650c7/healthbench_paper.pdf

[13] Khandekar, N. et al. Adv. Neural Inf. Process. Syst. 37 (NeurIPS 2024 Datasets and Bench-marks Track). doi:10.48550/arXiv.2406.12036.

[14] Hager, P. et al. Evaluation and mitigation of the limitations of large language models in clinical decision-making. Nat. Med. 30, 2613–2622 (2024). doi:10.1038/s41591-024-03097-1.

[15] Johri, S. et al. An evaluation framework for clinical use of large language models in patient interaction tasks. Nat. Med. 31, 77–86 (2025). doi:10.1038/s41591-024-03328-5.

[16] Wornow, M. et al. The shaky foundations of large language models and foundation models for electronic health records. npj Digit. Med. 6, 135 (2023). doi:10.1038/s41746-023-00879-8.

[17] Wilson, E. B. Probable inference, the law of succession, and statistical inference. J. Am. Stat. Assoc. 22, 209–212 (1927). doi:10.1080/01621459.1927.10502953.

